# Epigenetic mechanisms of osteoarthritis risk in human skeletal development

**DOI:** 10.1101/2024.05.05.24306832

**Authors:** Euan McDonnell, Sarah E Orr, Matthew J Barter, Danielle Rux, Abby Brumwell, Nicola Wrobel, Lee Murphy, Lynne M Overmann, Antony K Sorial, David A Young, Jamie Soul, Sarah J Rice

## Abstract

The epigenome, including the methylation of cytosine bases at CG dinucleotides, is intrinsically linked to transcriptional regulation. The tight regulation of gene expression during skeletal development is essential, with ∼1/500 individuals born with skeletal abnormalities. Furthermore, increasing evidence is emerging to link age-associated complex genetic musculoskeletal diseases, including osteoarthritis (OA), to developmental factors including joint shape. Multiple studies have shown a functional role for DNA methylation in the genetic mechanisms of OA risk using articular cartilage samples taken from aged patients. Despite this, our knowledge of temporal changes to the methylome during human cartilage development has been limited.

We quantified DNA methylation at ∼700,000 individual CpGs across the epigenome of developing human articular cartilage in 72 samples ranging from 7-21 post-conception weeks, a time period that includes cavitation of the developing knee joint. We identified significant changes in 8% of all CpGs, and >9400 developmental differentially methylated regions (dDMRs). The largest hypermethylated dDMRs mapped to transcriptional regulators of early skeletal patterning including *MEIS1* and *IRX1*. Conversely, the largest hypomethylated dDMRs mapped to genes encoding extracellular matrix proteins including *SPON2* and *TNXB* and were enriched in chondrocyte enhancers. Significant correlations were identified between the expression of these genes and methylation within the hypomethylated dDMRs. We further identified 811 CpGs at which significant dimorphism was present between the male and female samples, with the majority (68%) being hypermethylated in female samples.

Following imputation, we captured the genotype of these samples at >5 million variants and performed epigenome-wide methylation quantitative trait locus (mQTL) analysis. Colocalization analysis identified 26 loci at which genetic variants exhibited shared impacts upon methylation and OA genetic risk. This included loci which have been previously reported to harbour OA-mQTLs (including *GDF5* and *ALDH1A2*), yet the majority (73%) were novel (including those mapping to *CHST3, FGF1* and *TEAD1*).

To our knowledge, this is the first extensive study of DNA methylation across human articular cartilage development. We identify considerable methylomic plasticity within the development of knee cartilage and report active epigenomic mediators of OA risk operating in prenatal joint tissues.

## Introduction

The development of human limbs and synovial joints commences between four and eight weeks post conception (pcw). During limb bud outgrowth, mesenchymal condensations form the cartilage anlagen (driven by the transcriptional master-regulator, SOX9) to establish the rudimentary embryonic skeletal elements. The limb synovial joints are established through coordinated dedifferentiation of the nascent cartilage anlagen at the presumptive joint locations to form a unique pool of progenitors at each location known as the interzone. From these progenitors, all structures of the mature synovial joint are established including articular cartilage, synovial lining, meniscus and intrajoint ligaments. For the knee joint, chondrogenesis within the femoral condyles is advanced by the start of seven pcw (Carnegie Stage (CS) 19), and a clear interzone has formed between the femur and the tibia, with full cavitation of the joint and formation of the synovial cavity commencing at eight pcw^1^. The first appearance of menisci and ligaments also occurs within a similar developmental window, whilst the formation of the infrapatellar fat pad does not appear until much later in development (18pcw)^1^. This dynamic process of limb and synovial joint development requires the orchestrated expression genes essential for the development of defined and differentiated tissues^2^. The transcriptome is primarily regulated through the spatiotemporal expression of transcription factors (TFs), yet epigenetic processes, including DNA methylation (DNAm) both underlie and reinforce transitional plasticity during development^3^.

To date, DNAm remains the most extensively studied mammalian epigenetic mechanism. Methylation of DNA occurs at Cytosine-phosphate-Guanine (CpG) dinucleotides, of which there are approximately 28 million within the human genome. CpGs are recognised by DNA methyltransferase (DNMT) enzymes, which actively catalyse the addition of a methyl group from a S-adenosyI-L-methionine (SAM) donor to form 5-methyl cytosine (5mC)^4^. DNAm is intrinsically linked to transcriptional regulation, ostensibly to gene repression, by preventing binding of transcriptional activators to promoter regions, and further through the recruitment of repressive methyl-binding proteins. However, the relationship between DNAm and gene expression is far from straightforward, with gene body methylation often being associated with active transcription^5^. It is generally considered that DNAm within *cis-*regulatory elements (CREs) is repressive to the expression of the gene target^6,7^.

The methylome of human articular cartilage has been extensively studied, primarily in the context of the chronic joint disease osteoarthritis (OA). OA is a global leading cause of disability amongst older adults, hallmarked by the degradation of articular cartilage in the joints, most commonly the hip or knee. Studies of the aged cartilage methylome have revealed distinct epigenomic signatures between disease states^8,9^, and between joint sites themselves^10,11^, increasing our understanding of the joint specificity of disease. OA is multifactorial, with genetic risk factors contributing to ∼30% of the lifetime risk of developing knee OA. The integration of DNAm data into genetic studies, such as the statistical fine mapping of GWAS signals, has identified the co-localisation of methylation quantitative trait loci (mQTLs) with genetic risk signals^8,11–13^. This interplay between DNA sequence and CpG methylation status^14^ has further been shown to underpin tissue-specific molecular mechanisms of gene expression within the joint^15,16^.

Developmental factors also play a role in the risk of OA onset and progression in older age^17^. This includes both the shape of the joint^18–20^ (which affects the biomechanical properties and weight-bearing capacity) and the biochemical composition of the articular cartilage (which affects the resilience of the tissue to withstand stresses throughout the life course). Our recent targeted study of 39 CpGs investigated the presence of OA mQTLs in human foetal limbs, at seven well-characterised OA genetic risk loci^21^. We identified that at 85% of the CpGs, the significant OA-mQTLs replicated in the foetal tissues, demonstrating for the first time that functional epigenetic mechanisms associated with a musculoskeletal disease of older age operate from the start of life^21^.

Murine knockout or inactivation of the three known DNA methyltransferases (DNMTs) is lethal embryonically (*Dnmt1* and *Dnmt3b*) or postnatally (*Dnmt3a*), demonstrating the vital role of DNAm throughout development^22,23^. DNA methylating enzymes are expressed in proliferating chondrocytes during embryonic development and persist in articular chondrocytes during post-natal development^24^. Investigation of the methylome of human developmental cartilage has been limited to two studies to date, which utilised small sample numbers (n=8) falling within a narrow developmental window (14-19pcw)^25^, or, in the case of our previous study, applying targeted approaches which captured only a handful of CpGs^21^. Here, we present a comprehensive study of the methylomic trajectory of human articular cartilage in a broader window of development to capture novel risk loci for joint disease.

In this study, we examine the DNA methylome of 72 foetal cartilage samples from the developing knee (distal femur) between 7 (CS 23) and 21pcw. We identify the presence of differentially methylated regions across this developmental timeframe and examine their enrichment in CREs and TF binding sites. Furthermore, we investigate sexual dimorphism within the cartilage methylome during human development. Importantly, we identify the presence of developmental epigenomic changes contributing to the risk of OA in older life through the identification of foetal cartilage mQTLs co-localising with OA genetic risk signals.

## Methods

### Sample Collection

Cartilaginous tissue from the distal end of the developing human femur was supplied from the MRC and Wellcome Trust-funded Human Developmental Biology Resource (HDBR) at Newcastle University (http://www.hdbr.org, project number 200363), Newcastle upon Tyne. Tissues were obtained with appropriate maternal written consent and approval from the Newcastle and North Tyneside NHS Health Authority Joint Ethics Committee. HDBR is regulated by the UK Human Tissue Authority (HTA; www.hta.gov.uk) and operates by the relevant HTA Codes of Practice.

### Generation of methylation data and genotyping

DNA was genotyped using Illumina Global Screening Array v3 BeadChip at the Edinburgh Clinical Research Facility. Bisulphite conversion of DNA (500ng) was conducted using the EZ DNA methylation kit (Zymo) with Illumina-advised adjustments to the standard protocol. Methylation data was generated using the Illumina Methylation850 EPIC v1 array. The arrays were imaged on the Illumina iScan platform. Detected and autosomal probes were retained while those detected by less than 3 beads, with low detection P-values (P<0.001), present at known SNP sites, or known to be cross-reactive were removed from the analysis^26–28^.

### Transcript expression analysis measured by RT-qPCR

RNA (1μg) was reverse transcribed with the SuperScript IV cDNA synthesis kit (Invitrogen). The cDNA product was then used for quantitative real-time PCR (RT-qPCR) analysis following 1:20 dilution. Gene expression was quantified using TaqMan chemistry (QuantStudio3, Thermo Fisher). Pre-designed TaqMan assays (Integrated DNA Technologies, Belgium) were used to measure the expression of the target genes (Table S1). Gene expression was analysed relative to the expression of three housekeeping genes: *18S, GAPDH* and *HPRT1* using the 2^-Δct^ method as previously described^11^.

### mQTL analysis

mQTL analysis was performed using the matrixEQTL R package (v2.3), including Sentrix ID (batch), sex and gestation stage as covariates and testing for *cis*-associations based on a distance threshold of 500kb^29^. To account for ancestry, zero to four ancestry principal components (PC) from the 1000 genomes PC analysis (PCA) were trialled for inclusion in the mQTL analysis. The number of nominally significant mQTLs was compared between the different models and two PCs (79.86% of the total variance) were included in the final model. Resultant mQTLs were filtered based on an FDR < 0.05 or a conservative Bonferroni correction calculated using the matrixEQTL-reported total number of statistical tests. Significant mQTLs were associated with the protein-coding genes that they were closest to, based on EnsDb (v75) annotations^30^.

### Colocalisation analysis

The previously reported 100 independent genome-wide significant SNVs associated with 11 OA skeletal site phenotypes were used to assess colocalisation with the mQTL data^31^. Summary statistics for each phenotype were obtained from the musculoskeletal knowledge portal (https://msk.hugeamp.org/). MAF was calculated from the effect allele frequency, and case and control sizes for each phenotype were extracted from the respective publication. To assess mQTLs within a range of ±500kb from independent OA risk loci signal SNVs, we repeated the mQTL analysis while retaining all mQTL results (pvOutputThreshold=1). For methylation probes within ±500kb of the risk loci, we tested associated SNVs for colocalisation with genetic variants found in both the *cis* mQTL and the GWAS data. Colocalisation analyses were performed separately for each GWAS phenotype using the coloc R package^32^ (v 5.1.0) and the coloc.abf function, which calculates posterior probabilities for four hypotheses:

H0: Neither trait has a genetic association in the region.

H1: Only trait 1 has a genetic association in the region.

H2: Only trait 2 has a genetic association in the region.

H3: Both traits are associated but with different causal variants.

H4: Both traits are associated and share a single causal variant.

A posterior probability of H4 > 0.8 was considered as evidence for colocalisation. We further filtered results to retain SNVs present in our imputed genotype data and where the mQTLs was nominally significant (P<0.05).

## Results

### The DNA methylome remains dynamic throughout cartilage development

We isolated articular cartilage from the distal femur of 72 samples ranging from 7-21pcw (Fig.S1A). The mean(±SEM) gestational stage was 13.3±0.4 pcw (Fig.1A). Within the isolated cartilage, gene expression analysis of chondrocyte progenitor markers *GDF5* and *SOX9* showed a significant decrease in expression with increasing developmental stage (P=0.011 and 0.006, respectively; Fig.S1B-C). Conversely, the expression of articular chondrocyte marker *PRG4* significantly increased with stage (P=0.003; Fig.S1D). The Type II collagen gene, *COL2A1,* was highly expressed throughout the stages and did not significantly change (P=0.219; Fig.S1E).

**Figure 1.**
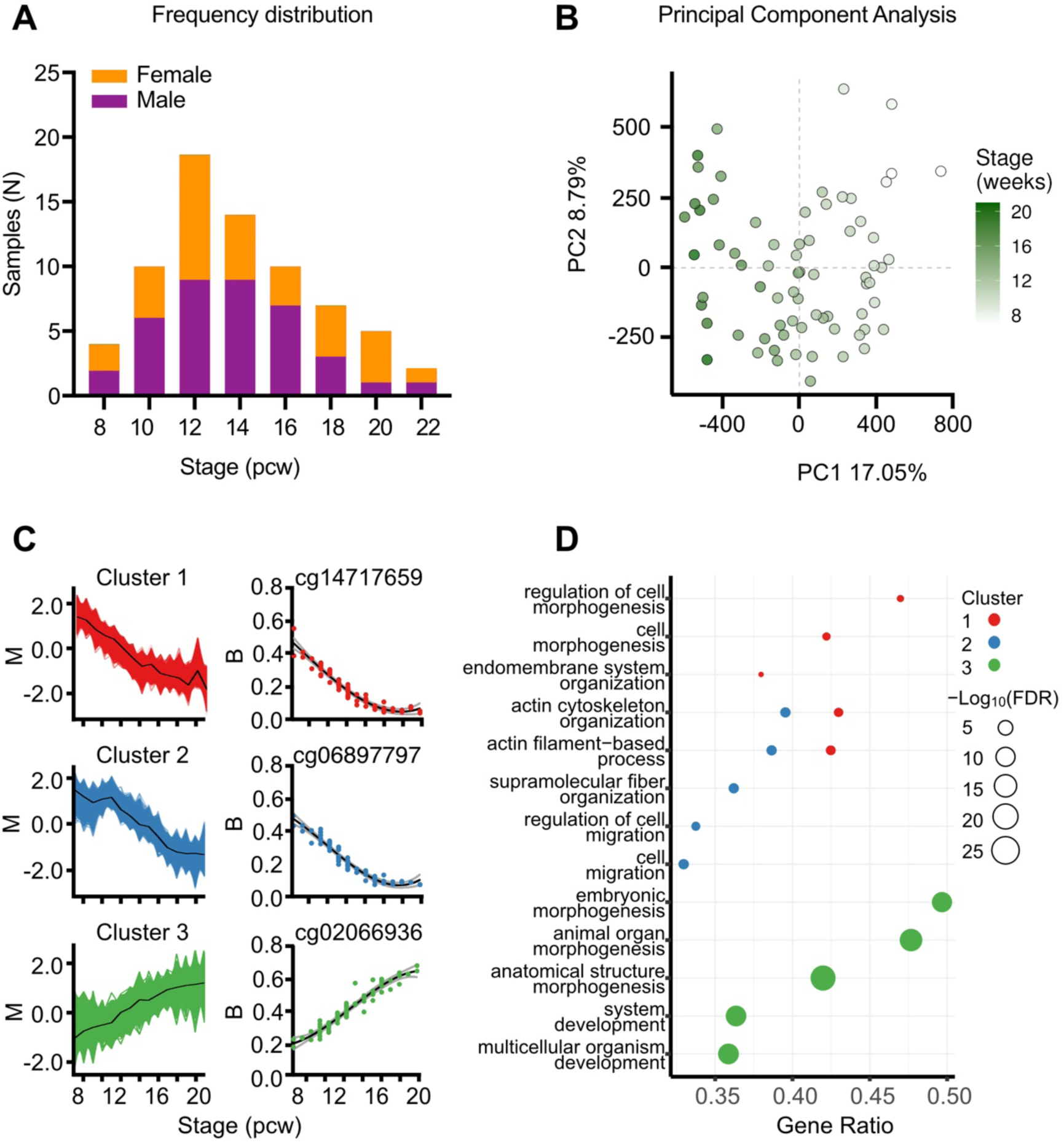
DNA methylation significantly changes at individual CpGs in foetal knee cartilage across the 7-21pcw developmental window. **A,** Histogram of samples (N=71) showing the distribution of developmental stage. Purple, Male; Orange, female. **B,** Principal component analysis of the samples revealed that PC1 is associated with the developmental stage. **C,** The DMPs fell into three distinct clusters based upon the methylation trends across the developmental window. The mfuzz plot for each cluster is displayed, along with the most significantly differentially methylated CpG belonging to each cluster. **D,** Gene ontology (GO) analysis of the top significant biological processes enriched in each of the three clusters. Gene ratio is the ratio of the proportion of genes in a GO term annotated with at least one significant CpG relative to the proportion of genes in a GO term annotated with a detected CpG.

We profiled the DNA methylation (DNAm) of the 72 samples using Illumina HumanMethylation850 EPIC v1 microarrays. One sample was dropped due to mismatch between predicted and labelled sex. The remaining 71 samples (Table S2) had a minimum bisulphite conversion rate of 89.69% and following QC we retained data at 678,267 CpGs. Data were normalised via quantile-normalisation, followed by principal component analysis (PCA), where developmental stage was found to strongly correlate with PC1, which accounted for 17% of the total variance in global methylation (Fig. 1B). Across the captured developmental window, we identified 53,800 significantly differentially methylated probes (DMPs; FDR<0.001, log2fc>0.1; Table S3), with 48% becoming significantly hypomethylated through skeletogenesis. We analysed the trend in methylation changes across the developmental stages, identifying three clusters. Clusters 1 and 2 became significantly hypomethylated across the developmental window (Fig. 1C; red and blue, respectively). The primary difference separating the two groups was an observable lag in decreased methylation in Cluster 2, where DNAm remained stable until >10pcw. Conversely, the CpGs belonging to Cluster 3 (Fig. 1C; green) became hypermethylated throughout development. GO analysis revealed similar and shared significant terms (FDR<0.05) for Clusters 1 and 2, including biological processes such as “actin filament-based processes” and “cell morphogenesis” (Fig.1D) mapping to cellular compartments including “actin cytoskeleton” and “focal adhesion” (Table S4). Analysis of Cluster 3 identified significant enrichment of biological processes including “embryonic morphogenesis” and “anatomical structure development” (Fig.1D) and the cellular compartments “collagen-containing extracellular matrix” and “transcriptional regulator complex” (Table S4).

To gain further insight into the biological role of the DMPs, we next investigated the presence of differentially methylated regions (DMRs) based upon regions of DNAm co-regulation. We identified 9,462 developmental DMRs (dDMRs) ranging in size between 2 and 59 CpGs (mean = 2.9) consisting of 27,447 individual sites (FDR<0.001, log2fc>0.1; Fig.2A, Table S5). In Figure 2A the most significant dDMRs (irrespective of size) are annotated with the nearest protein coding gene. The largest hypermethylated dDMRs (n=21-59) mapped to genes encoding known transcriptional regulators of early development including the *HOXA* gene locus and *TBX3* (Fig.2B). GO analysis revealed 326 significant (FDR<0.01) biological processes associated with hypermethylated dDMRs (Table S6). The top 20 most significant biological process terms included “skeletal system development”, “anatomical structure development”, and “embryonic morphogenesis” (Fig.2C). Just 20 biological processes were significantly associated with hypomethylated regions (FDR<0.01; Table S7), which mapped to genes encoding ECM proteins including *TNXB* (Tenascin-X) and *SPON2* (Spondin-2; Fig.2B0. “Supramolecular fiber organisation” was the highest enriched biological process (gene ratio, 0.22; FDR = 0.006) along with multiple terms relating to the innate immune system (Fig.2C).

**Figure 2.**
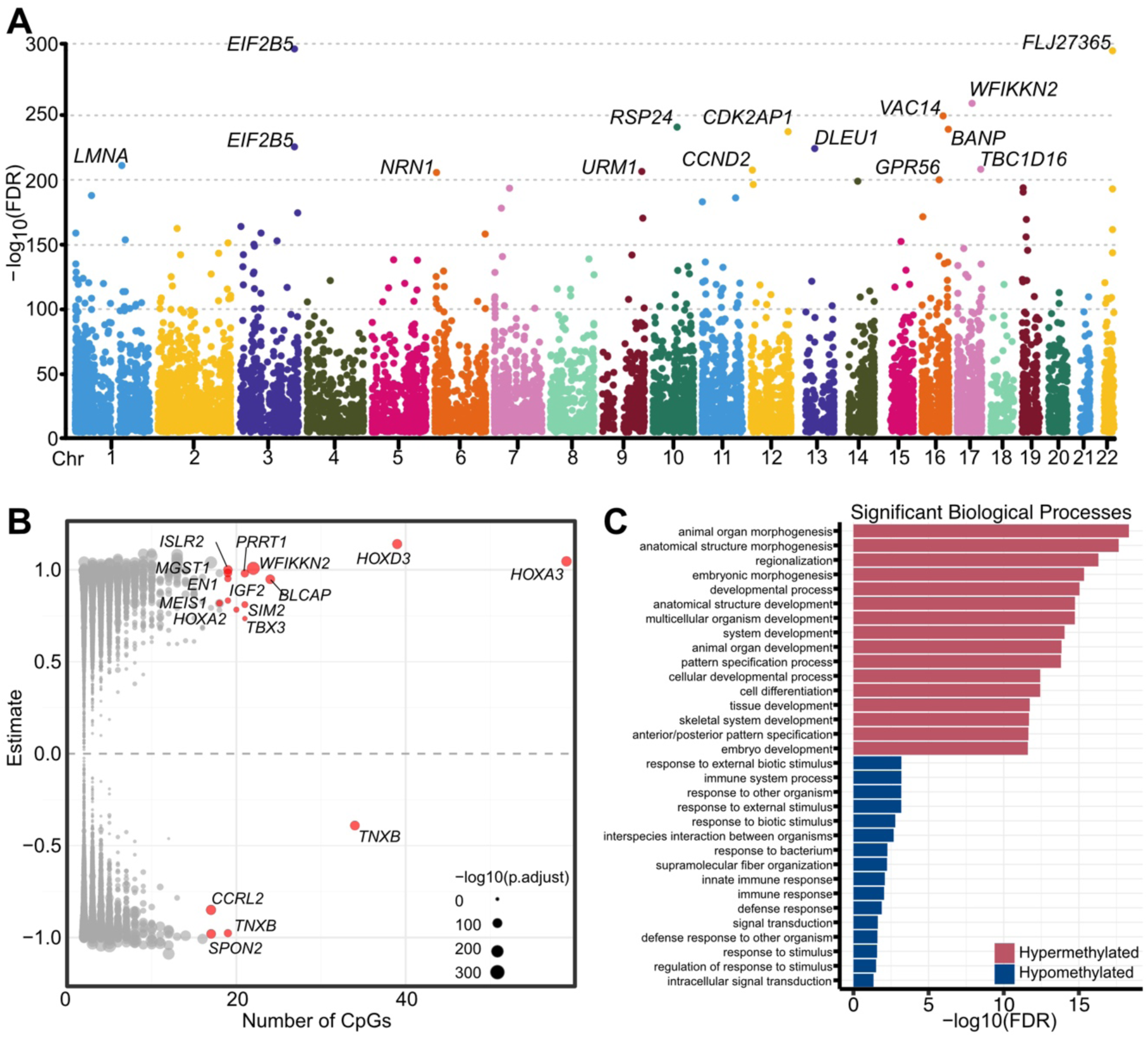
Co-regulation of DNAm at individual CpGs results in developmental differentially methylated regions (dDMRs) in foetal knee cartilage. **A,** Manhattan plot of dDMRs across the epigenome. The nearest protein-coding gene to the most significant dDMRs is labelled. **B,** Volcano plot of dDMRs by size (number of CpGs). The most significant are highlighted in red with the nearest protein-coding gene labelled. **C,** Gene ontology (GO) analysis of the most significant biological processes enriched in hypermethylated (red) and hypomethylated (blue) dDMRs.

### Developmental DMRs overlap with open chromatin regions and correlate with gene expression

We next intersected the physical location of the dDMRs with open chromatin peaks from 12pcw foetal distal femoral cartilage. We identified overlap between 3792 dDMRs (40%), mapping to 2558 genes, further highlighting those most likely to impart regulatory function during development (Table S8). The largest hyper- and hypomethylated regions overlapping with foetal knee ATAC peaks are displayed in Table 1.

**Table 1.**
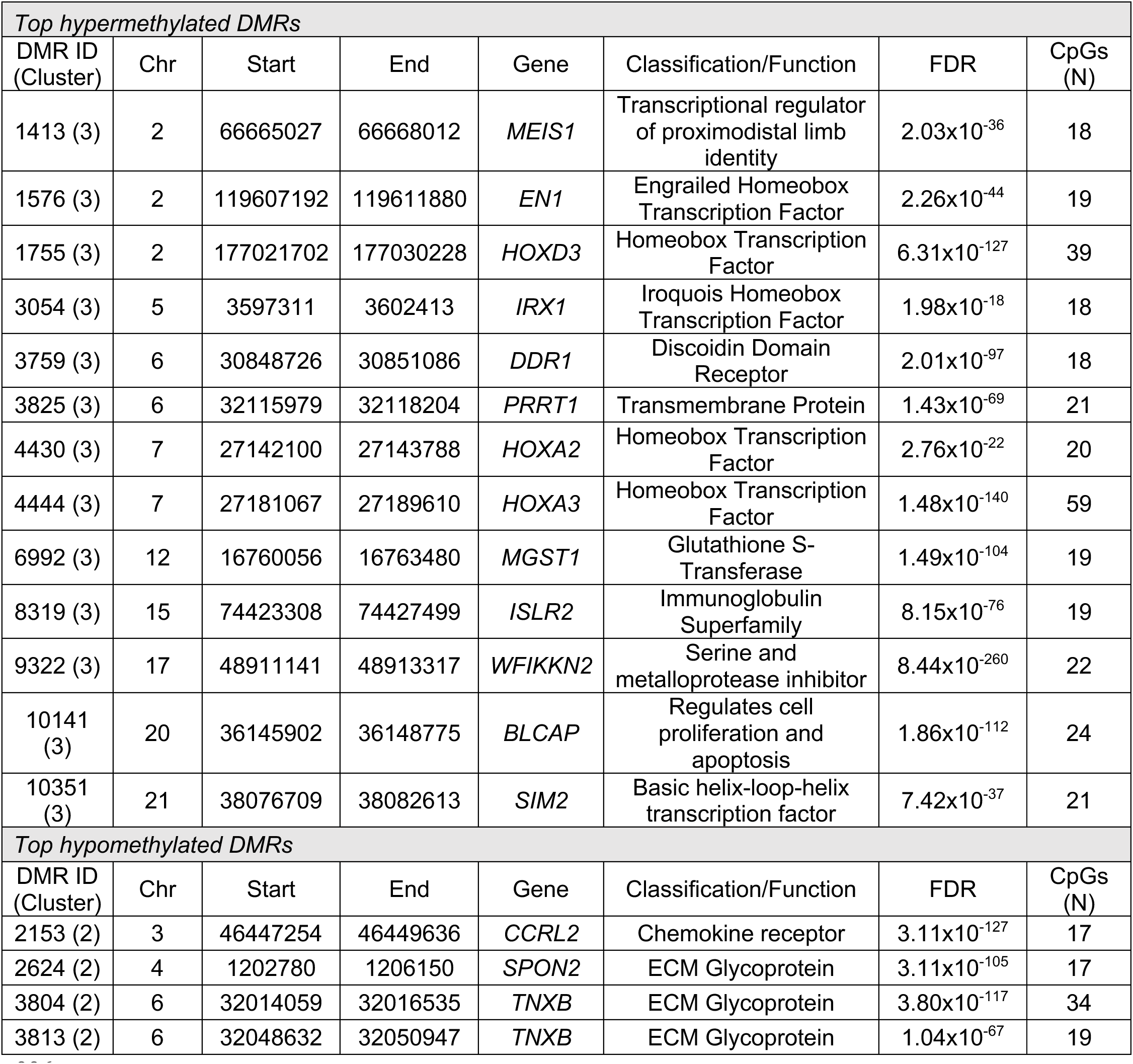
Top Developmental DMRs which intersect with open chromatin regions in developing knee cartilage. The largest identified dDMRs (N CpGs>16) which became either hypermethylated (upper) or hypomethylated (lower) across the captured developmental window. The nearest physical gene to each dDMR is listed.

To investigate the putative functional role of these DMRs in developmental gene regulation, we measured the expression eight of the genes mapping to the dDMRs presented in Table 1. The six transcripts mapping to hypermethylated regions (*MEIS1, EN1, HOXD3, IRX1, HOXA3, SIM2*) were expressed at low levels throughout the captured developmental window (Fig.S2, red). The expression of *HOXA3* negatively correlated with DNAm at 25/59 individual CpGs within the dDMR (P=0.048-0.008; Table S9, Fig.3A LHS). Interestingly, the *HOX* genes mapping to this dDMR do not have reported functions in limb development. We compared their expression to that of *HOXA10* and *HOXA11*, which have recently been reported to be expressed specifically in the zone between the proximal and distal human hindlimb at 5.6pcw, in the region where the knee cavity will form^33^. These genes are also located at the *HOXA* gene locus, a region of tightly controlled co-regulation (Fig.3A upper panel). However, their expression showed no correlation with DNAm within this region (P>0.05; Fig.3A middle and RHS; Table S9).

**Figure 3.**
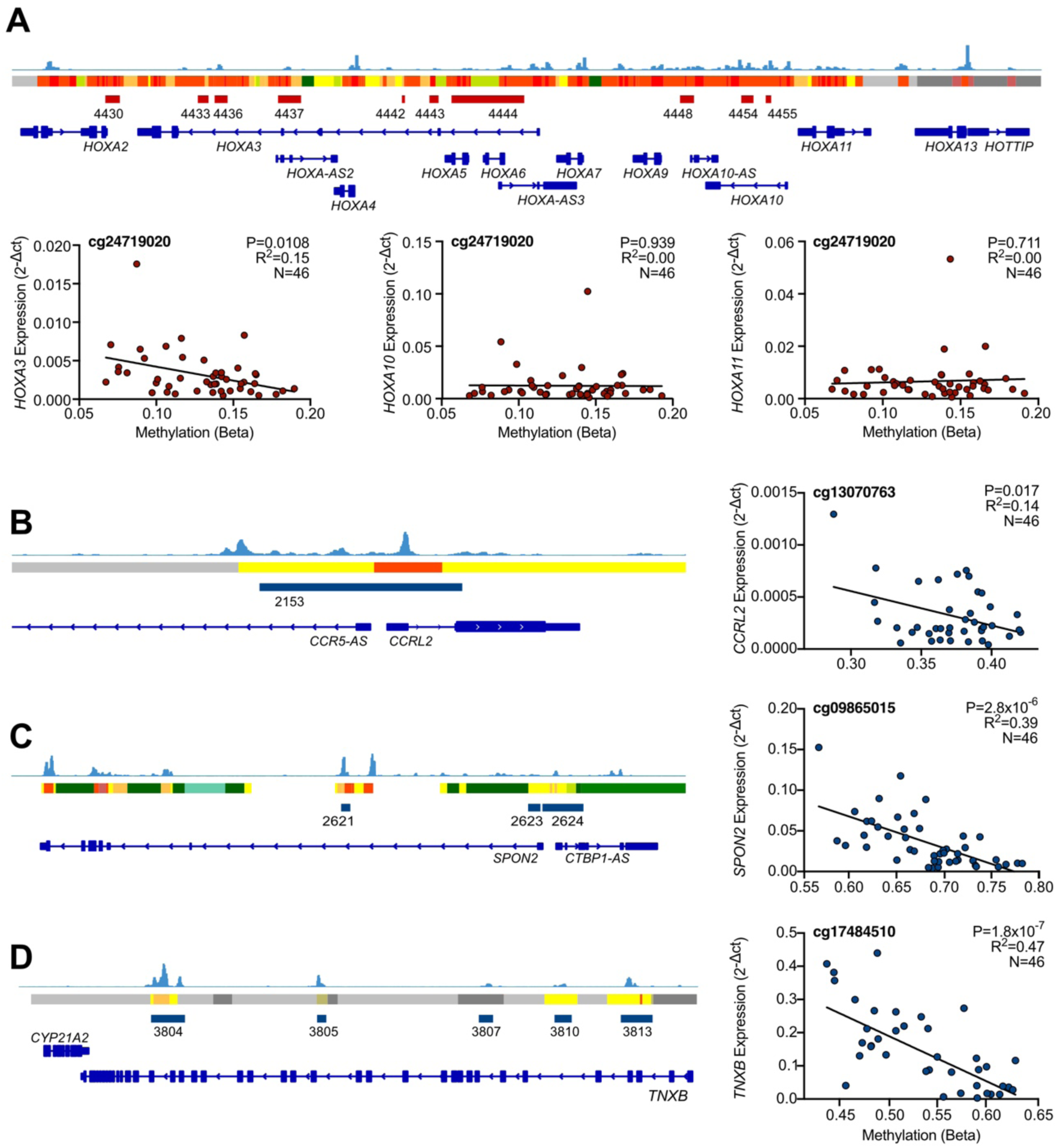
The expression of genes mapping to dDMRs significantly correlates with methylation within the regions. **A,** (Upper) Screenshot of the Integrated Genome Viewer (IGV) displaying the HOXA gene locus on chromosome 7. Foetal knee cartilage ATAC-seq peaks (light blue) show regions of open chromatin. Below the peaks, the ROADMAP chromatin state track from human cultured chondrocytes (E049) is displayed. Red, active transcription start site; yellow/orange, enhancer; green, transcribed; grey, repressed. Hypermethylated dDMRs are shown in dark red and annotated with the dDMR ID. Genes within the region are shown in bright blue with the gene names labelled. (Lower) Gene expression of HOXA3 (LHS), HOXA10 (middle) and HOXA11 (RHS) measured by RT-qPCR regressed against DNAm at cg24719020, which falls within dDMR 4444. **B-D, (LHS)** IGV screenshots of chromosomes 3, 4, and 6, respectively. Foetal ATAC-seq data is displayed as described in (A). Hypomethylated dDMRs are shown in blue. **RHS,** gene expression of CCRL2, SPON2, and TNXB regressed against DNAm at a single CpG within each of the dDMRs mapping to the respective genes. Statistical analysis was performed by multiple linear regression using methylation M-values. Beta values are displayed for ease of interpretation.

All three genes mapping to the largest hypomethylated regions, *CCRL2* (labelled dDMR 2153), *SPON2* (dDMR 2624) and *TNXB* (dDMRs 3804 and 3813), fell within putative chondrocyte enhancers and regions of open chromatin in foetal chondrocytes (Fig.3B-D). The expression of all three genes significantly increased in correlation with decreasing methylation within the mapped dDMRs at 3/17 (P=0.017 - 0.041), 6/17 (P=2.8x10^-6^ - 0.002) and 42/53 (P=1.82x10-^7^ – 0.017) CpGs, respectively (Fig.3B-D).

### Developmental DMRs are enriched in active enhancers and transcription factor binding sites

We performed motif enrichment analysis on the hyper- and hypomethylated dDMRs for enrichment of known transcription factor (TF) binding motifs. We identified significant enrichment (FDR≤0.05) of 48 known TF motifs on the HOCOMOCO database (Fig.4A, Table S10). Notably, the regions that decreased in methylation across the developmental window were enriched for TFs with known roles in chondrocyte biology and synovial joint formation including NFATC1 (P.adj=0.02)^34^, CREB5 (P.adj=9.7x10^-5^)^35^, and the FOS/JUN families of TFs (P.adj<8.1x10^-19^). The regions that increased in methylation across development were enriched for motifs of early transcriptional regulators of skeletogenesis including HOXD3 (P.adj=0.01) and CUX1/2 (P.adj<0.04; Fig.4A)^36^.

**Figure 4.**
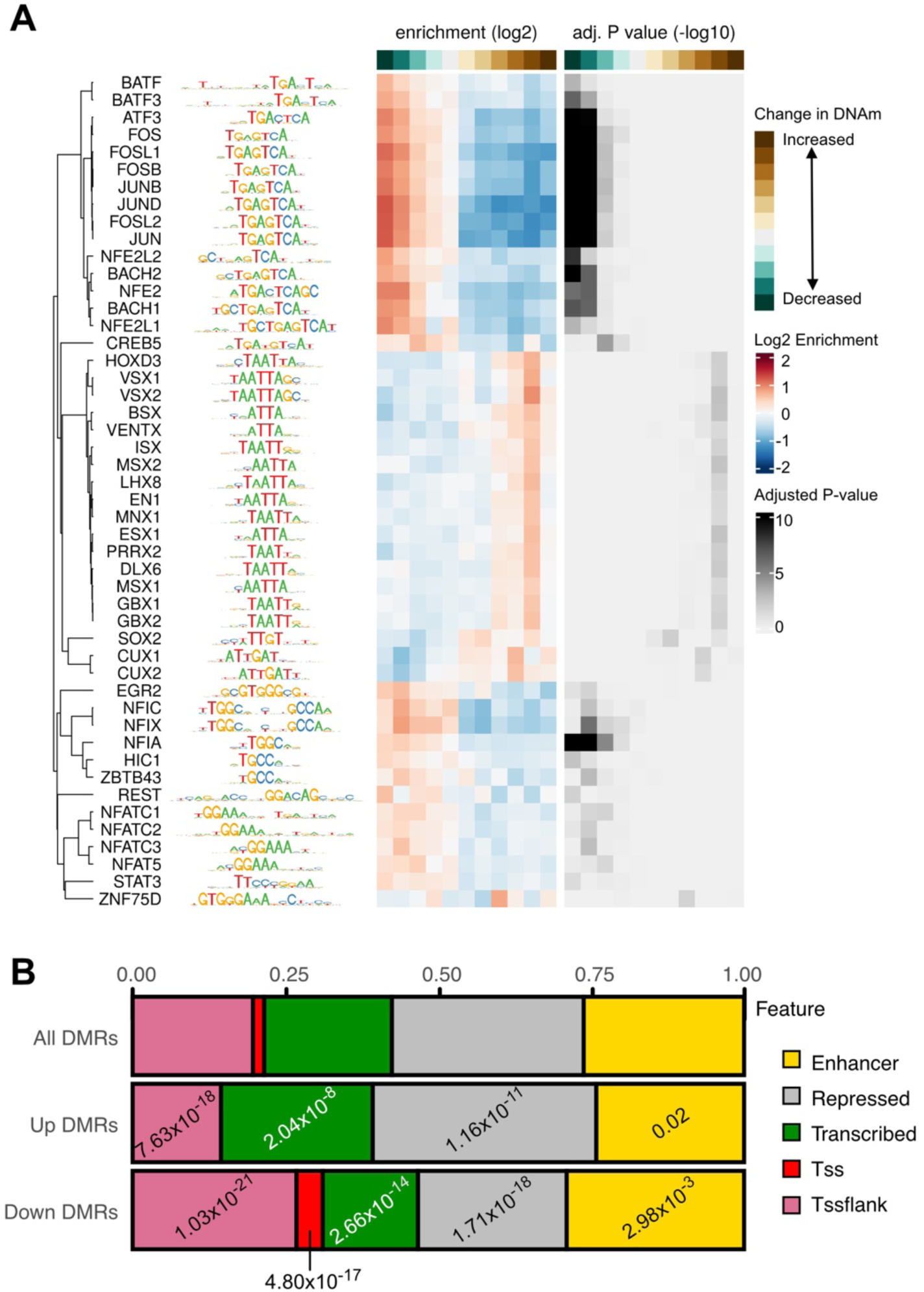
Developmental DMRs are enriched in regulatory elements. **A,** Transcription factor (TF) motifs taken from the Hocomoco database which are enriched in dDMRs **Left,** name and logo plot of enriched TF motif within the dDMRs, **Centre,** Heatmap of Log2 enrichment value of the motifs within hypermethylated (brown) or hypomethylated (green) dDMRs. **Right,** Heatmap of -Log10 adjusted P-value. Darker colour indicates higher statistical significance. **B,** Frequency of identified dDMRs overlapping with human cultured chondrocyte (Roadmap E049) chromatin states. Enhancer, yellow; repressed region, grey; transcribed region, green; transcription start site (TSS), red; region flanking TSS, pink. Statistical significance was calculated using Fisher’s exact test with Bonferroni correction with all significant DMRs as background. Significant under- or overrepresentation is labelled with the adjusted P-value.

Finally, we overlapped the dDMRs with Roadmap chromatin state data generated in human chondrocytes (E049) to test for overrepresentation amongst distinct categories of CREs. Hypermethylated dDMRs were significantly overrepresented in transcribed (Fig.4B, green; P.adj=2.0x10^-8^) and repressed regions (Fig.4B, grey; P.adj=1.2x10^-11^) when compared to all identified significant dDMRs. No hypermethylated dDMRs overlapped directly with transcriptional start sites (TSS; Fig.4B, red). Conversely, hypomethylated dDMRs were overrepresented in enhancers (P.adj=2.9x10^-3^) and TSS (P.adj=4.8x10^-17^) but underrepresented in repressed regions (P.adj=1.7x10^-18^), indicating that across development, regions becoming hypomethylated were predominantly regulating the function of CREs.

### Chondrocyte DNA methylation in articular cartilage development exhibits sexual dimorphism

Previously, methylome analysis performed in 358 blood samples from extremely premature neonates (at 23-27 weeks’ gestation, which relates to ∼21-25pcw) showed that a large proportion of CpGs exhibited sexually dimorphic levels of DNAm^37^. Using the EPIC array, Santos *et al.,* identified 5595 DMPs (FDR<0.00001), 95% of which were hypermethylated in females. We tested for significant sex-specific differences in DNAm amongst our developmental cartilage samples. Epigenome-wide analysis revealed 811 sex-specific DMPs (sDMPs; FDR<0.05), mapping to 534 genes (Table S11, Fig.S3A). Of these sites, 523 (64.5%) were hypermethylated in the female samples. Fifty-seven percent (461) of the sDMPs we identified in developing articular cartilage also showed significant sex effects in the neonatal blood dataset (Fig.S3B). There was a direct overlap between all of the top 20 reported probes, with methylation effects occurring in the same direction (Table 2, Fig.S3C). Amongst the genes mapping to these CpGs are *NAB1, RFTN1,* and *CEP170*.

**Table 2.** Top 20 sDMPs in developing articular cartilage. The most significant DMPs between male and female cartilage samples are listed. Chr, chromosome; FDR, false discovery rate; B, beta value; M, male; F, female.

| CpG ID | Chr | Position | FDR-adjusted P-value | Nearest Gene Symbol | Distance (bp) | Cartilage highest mean B | Blood highest mean B |
| --- | --- | --- | --- | --- | --- | --- | --- |
| cg03618918 | 1 | 160865097 | $1.59 \times 10^{-21}$ | *ITLN1* | 10136 | M | M |
| cg26919182 | 1 | 202522232 | $2.41 \times 10^{-22}$ | *PPP1R12B* | 0 | F | F |
| cg15817705 | 1 | 209406063 | $2.86 \times 10^{-23}$ | *CAMK1G* | 350998 | M | M |
| cg25742246 | 1 | 214153460 | $2.22 \times 10^{-19}$ | *PROX1* | 3063 | F | F |
| cg12691488 | 1 | 243053673 | $1.27 \times 10^{-39}$ | *CEP170* | 234056 | M | M |
| cg15228509 | 1 | 243074717 | $2.12 \times 10^{-42}$ | *CEP170* | 213012 | F | F |
| cg02989351 | 2 | 9770584 | $1.16 \times 10^{-23}$ | *YWHAQ* | 0 | F | F |
| cg03226871 | 2 | 128128958 | $2.88 \times 10^{-20}$ | *MAP3K2* | 0 | M | M |
| cg19765154 | 2 | 191524409 | $7.70 \times 10^{-36}$ | *NAB1* | 0 | M | M |
| cg20262915 | 2 | 191524489 | $2.02 \times 10^{-22}$ | *NAB1* | 0 | M | M |
| cg00148935 | 3 | 16398839 | $1.19 \times 10^{-27}$ | *RFTN1* | 0 | F | F |
| cg07850329 | 9 | 33265029 | $3.04 \times 10^{-21}$ | *CHMP5* | 0 | F | F |
| cg07852945 | 9 | 84303915 | $9.64 \times 10^{-20}$ | *TLE1* | 0 | F | F |
| cg02716779 | 9 | 131016143 | $3.27 \times 10^{-22}$ | *DNM1* | 0 | F | F |
| cg26355737 | 13 | 114292172 | $2.84 \times 10^{-21}$ | *TFDP1* | 0 | M | M |
| cg11284736 | 15 | 83826708 | $3.77 \times 10^{-21}$ | *HDGFRP3* | 0 | M | M |
| cg03626220 | 16 | 71052200 | $1.47 \times 10^{-24}$ | *HYDIN* | 0 | M | M |
| cg03218192 | 17 | 33914403 | $4.68 \times 10^{-22}$ | *AP2B1* | 0 | F | F |
| cg12607525 | 17 | 42286849 | $1.94 \times 10^{-22}$ | *UBTF* | 0 | M | M |
| cg17612569 | 21 | 27107221 | $2.38 \times 10^{-25}$ | *ATP5J* | 0 | M | M |

We further identified 60 significant sDMRs (FDR<0.01), consisting of 178 CpGs (Table S12). At 46 identified regions (77%), DNAm was significantly higher in the female samples than in male. No significant GO terms (all FDR>0.05) were associated with either the sDMPs or sDMRs, consistent with the findings of Santos *et al.,* in their analysis of neonatal blood samples^37^.

### mQTLs co-localizing with OA risk signals (OA-mQTLs) are active during articular cartilage development

We next investigated the presence of mQTLs in the developing human knee cartilage. Epigenome-wide analysis revealed 399,952 significant (Bonferroni-corrected P<0.05) *cis*-mQTLs (Table S13). The signals were widespread across the epigenome (Fig.5A) and consisted of 11,299 individual CpGs and 257,908 SNVs. The most significant mQTLs mapped to the genes *ELMOD1* (P.adj=3.7x10^-50^), *ATPIF1* (P=1.4x10^-48^), and *PHGDH* (P.adj=4.0x10^-47^).

**Figure 5.**
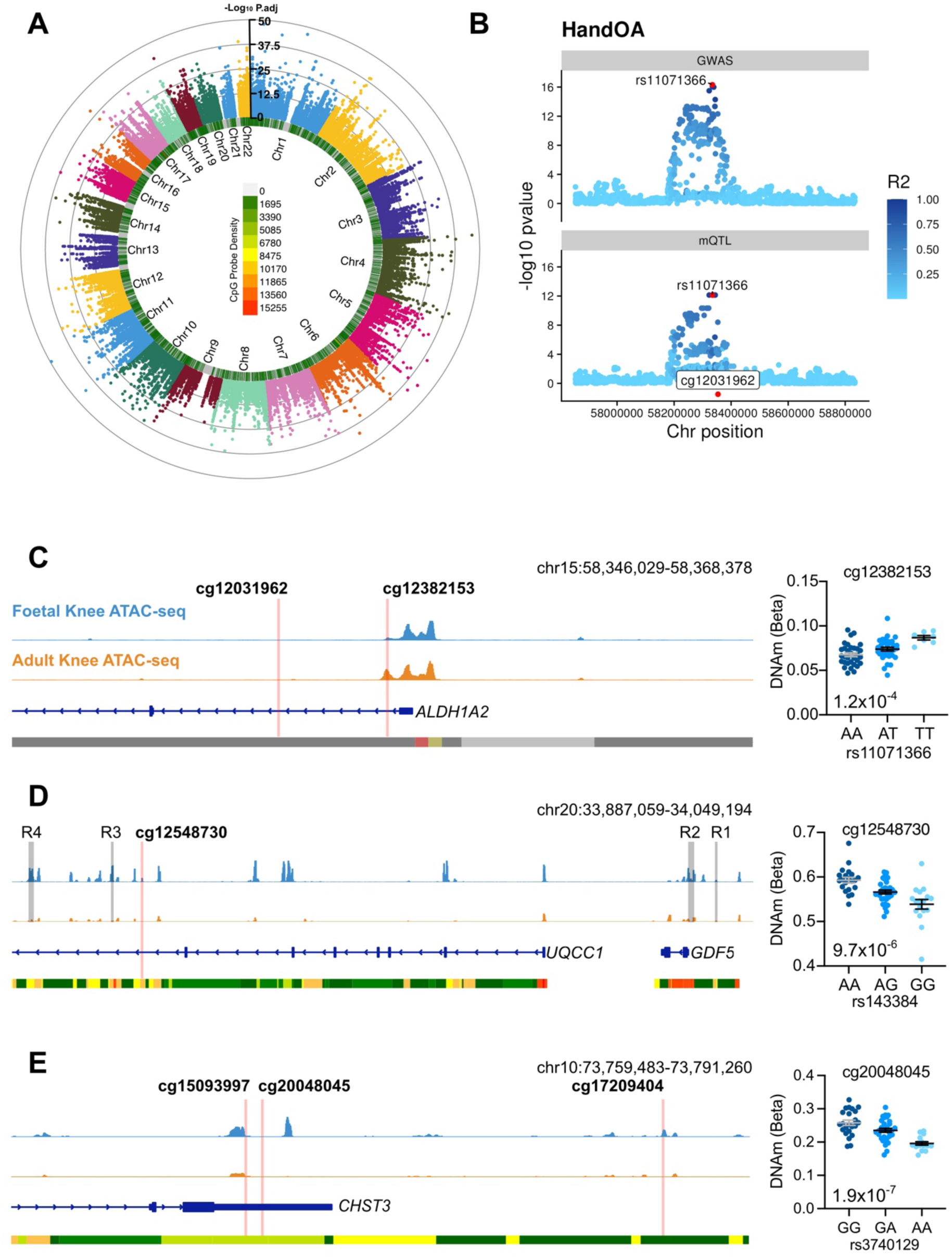
Epigenome-wide mQTL analysis reveals co-localisation with OA genetic risk signals in developmental cartilage. **A,** Manhattan plot of significant (Bonferroni adjusted P-value <0.05) epigenome-wide mQTL signals. The inner heatmap represents the Illumina probe density in the respective regions across the genome. **B,** An example of a colocalisation event. The mQTL for the methylation site cg02900766 (bottom) colocalised with the GWAS signal rs11071366 for hand OA in the same genomic region (top). Here, we observed a posterior probability (PP) for a shared causal variant of 99%. **C-E,** Integrated Genome Viewer (IGV) screenshot of the genetic region mapping to foetal cartilage mQTLs colocalising with OA GWAS signals rs11071366 (C), rs143384 (D), and rs3740129 (E). Light blue, foetal knee ATAC-seq data with peaks representing open chromatin regions; orange, OA knee ATAC-seq data; dark blue, gene transcript; bottom track is Roadmap chromatin state data from E049 cultured chondrocytes. Red, active transcription start site; yellow/orange, enhancer; green, transcribed; grey, repressed.

To identify shared causal impacts of SNV genotype upon OA risk and DNAm, we performed a colocalisation analysis of 100 risk signals from 11 OA phenotypes with the foetal cartilage mQTLs. In total, we identified 104 colocalisations with foetal cartilage mQTLs (posterior probability (PP) >0.8; Table S14) amongst 49 CpGs spanning 26 genomic loci (Table 3). At 7 of the loci, OA-mQTLs were identified which have previously been reported in investigations of aged adult cartilage. We overlapped the physical location of the CpGs with open chromatin regions in foetal and OA knee chondrocytes^21^, regions of H3K27ac enrichment in foetal limb tissues^38^, and with designated chromatin states in mesenchymal stem cells (MSCs) and chondrocytes^39^. Through this, we compiled *in silico* evidence of functional impact for the identified mQTLs restricted to development (D) or throughout the life course (L). Evidence supporting function was identified at 19 loci, with around half (9) consisting of mQTLs which fell within open chromatin or H3K27ac-enriched regions solely in developmental samples.

**Table 3.**
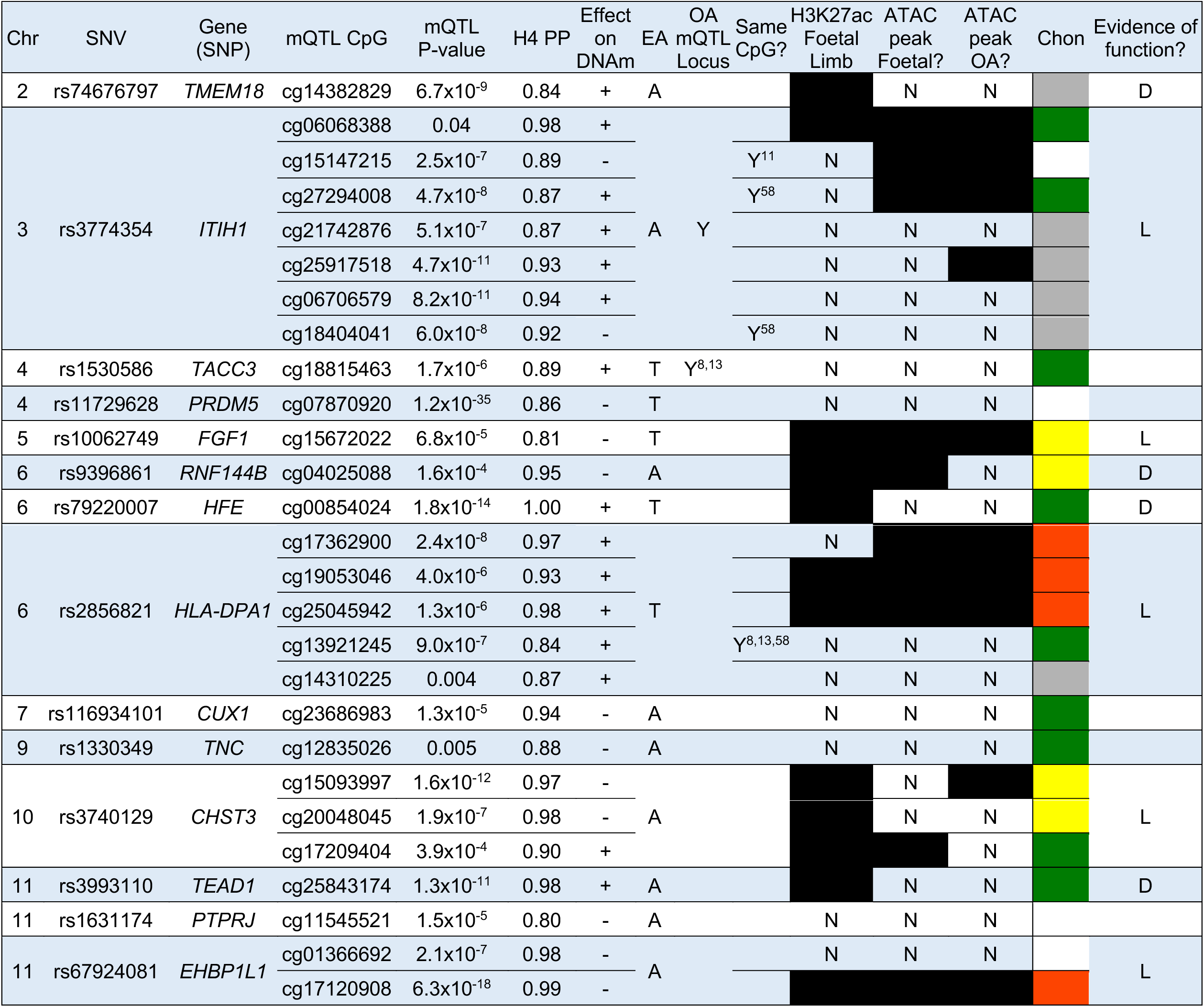
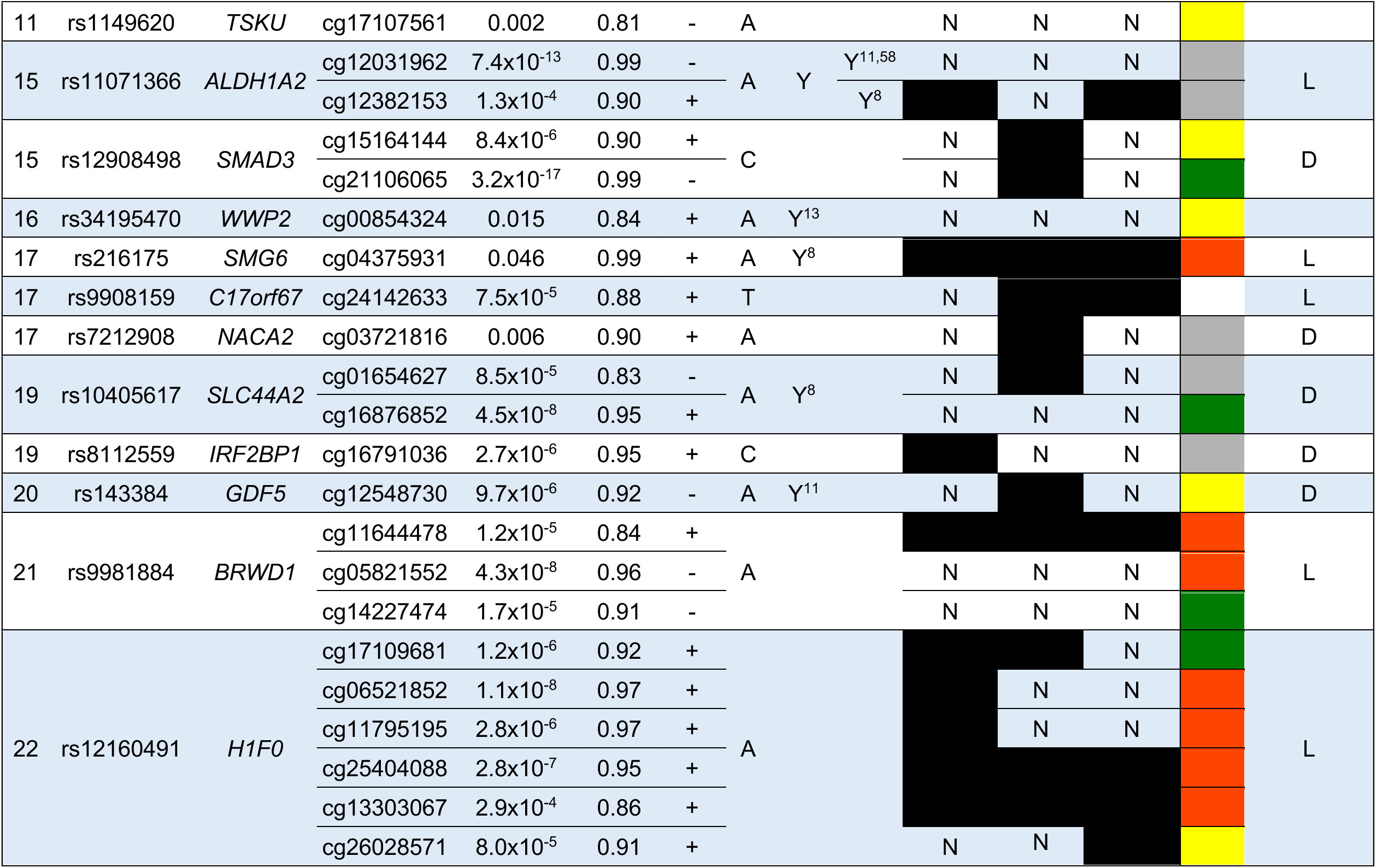
OA genetic risk loci co-localizing with significant mQTLs in human foetal knee cartilage. The nearest gene to the SNV is listed. MA, minor allele; EA, effect allele H4 PP, poster probability that both traits are associated and share a single causal variant. We investigated whether the CpGs fell within or adjacent to (within 150bp of) narrow peaks in our cartilage ATAC-seq data sets (GSE214394) or within regions enriched for H3K27ac (a marker of enhancer activity) in human limb tissues at embryonic day 47 (CS19; Gene Expression Omnibus accession GSE42413). In human cultured chondrocytes (E049) the 15 ROADMAP states were collapsed into 5 functional classifications: Yellow: enhancer (Enh, EnhG, EnhBiv), White: repressed (ZNF/Rpts, Quies), Grey: heterochromatic (ReprPC, ReprPCWk), Red: promoters (TssBiv, TssA, TssAFlnk, BivFlnk) and Green: transcribed (TxWk, Tx, TxFlnk). Regions were overlapped with the CpG locations and are indicated by the denoted colours. L, lifecourse; D, development

One example of OA-mQTLs operating in both foetal and aged human cartilage is on chromosome 15, where mQTLs at cg12031962 and cg12382153 map to *ALDH1A2* (P=7.4x10^-13^ and 1.3x10^-4^, respectively). These mQTLs co-localise with the primary phenotypes “Hand OA” (PP=0.99, Fig. 5B), “Thumb OA” (PP=0.99), “Total Knee Replacement” (PP=0.98) and “Finger OA” (PP=0.93; Table S14) marked by the risk SNV rs11071366 (A>T). At both CpGs, OA-mQTLs have previously been reported in adult cartilage (Table 3). The methylation site cg12382153 falls within an open chromatin peak flanking the *ALDH1A2* transcriptional start site (Fig.5C). Here, the major and OA effect allele, A, correlates with a decrease in DNAm at both CpGs, consistent with reports in aged cartilage^8^.

Conversely, on chromosome 20, a single intronic mQTL was identified which maps to a previously unreported site at a known OA risk locus, mapping to the gene *GDF5* (Fig.5D). Here, DNAm at cg12548730 co-localised with the OA risk SNV rs143384 in the “All OA” (PP = 0.916), “Total Joint Replacement” (PP = 0.915) and “Knee and Hip OA” (PP = 0.891) primary phenotypes (Table S14). The CpG is located within an intron of *UQCC1*, downstream of previously reported *GDF5* cartilage enhancers R3 and R4^40,41^. The OA effect allele (G) significantly correlated with lower levels of DNAm (P=9.7x10^-6^; Fig.5D). Analysis of our existing knee cartilage ATAC-seq dataset in human foetal (distal femur) and adult cartilage samples showed that the CpG is located adjacent to an open chromatin peak in foetal cartilage (blue), which is not an accessible region in OA cartilage (orange). This region has not previously been investigated as a regulatory element for *GDF5.* A previous mQTL at this locus has been reported in adult OA cartilage, located adjacent to the promoter of *UQCC1* (cg14752227)^11^.

At the majority of the loci (19), no previous reports of OA-mQTLs have been reported through analyses of adult cartilage. On chromosome 10, three OA-mQTLs were identified that fell within putative regulatory elements of *CHST3,* encoding carbohydrate sulfotransferase 3 (Fig.5E). Two of the CpGs, cg15093997 and cg20048045, located within the 3’UTR of the gene, near to open chromatin regions (Fig.5E). At both CpGs, the effect allele, A, at rs3740129 (G>A) correlated with significantly decreased levels of DNAm (P=1.6x10^-12^ and 1.9x10^-7^, respectively). Conversely, at the more distal CpG (cg17209404), located within an intergenic open chromatin region only in foetal cartilage, DNAm was significantly increased in the presence of the OA effect allele (P=3.9x10-4; Table S14).

### Genetic and epigenetic effects influence distinct subsets of CpGs comprising the foetal cartilage methylome

Finally, we tested whether the 11,299 CpGs which comprise the identified significant mQTLs across the epigenome are amenable to external influences occurring between sexes (811 sDMPs) and through articular cartilage development (80,536 dDMPs). Between the mQTLs and dDMPs, an overlap of just 744 CpGs was identified, indicating that only 6.6% of CpGs that are significantly regulated by DNA sequence are impacted by developmental stage (P=5.2x10^-80^). Similarly, between the mQTL CpGs and the 811 sDMPs, an overlap of only 6 CpGs (0.74%) was observed (P= 0.0184). This indicates that the CpGs at which DNAm is heavily influenced by the genetic architecture of the region are less amenable to environmental influences on the epigenome.

## Discussion

In this study, we present the epigenomic trajectory across the development of human articular cartilage. To our knowledge, this provides the first insight into methylomic plasticity across a large window of human skeletogenesis. We identified that developmental stage is the predominant determinant of methylation status within our samples, with ∼8% of CpGs significantly changing across the captured timeframe. We further identified >9400 dDMRs, 40% of which intersected with open chromatin regions in foetal knee cartilage (12pcw), providing evidence for the potential functionality of the regions. We demonstrated that 811 of the captured CpGs exhibited sexually dimorphic effects, with the majority (64%) being hypermethylated in female samples. Finally, we conducted an epigenome-wide mQTL analysis and found strong evidence of co-localizations of these molecular phenotypes with OA GWAS signals, building upon existing evidence of developmental origins of a complex genetic disease of older age.

In 2023, Zhang *et al* published the first single-cell and spatial transcriptomic atlas of the developing human hindlimb^33^. They provided evidence of nascent articular chondrocytes that expressed high levels of *PRG4* in the distal femur by 8pcw. In our data, the overwhelming impact of the developmental stage upon the cartilage methylome partly reflects the changing nature of the cartilage cellular composition captured in our bulk analysis. In earlier stages (<12pcw) we have likely captured epiphyseal chondrocytes along with the nascent articular cartilage that will continue to develop postnatally, whereas after 12pcw, we were able to isolate the nascent articular cartilage more specifically. Our gene expression profiling data provide strong evidence that the primary population of cells analysed in this study are nascent epiphyseal and articular chondrocytes. Underscoring this, our data also align with studies of embryonic development of epiphyseal and articular cartilage in mouse models where time-course expression and lineage tracing studies have shown that both *Gdf5* and *Sox9* expression decreases, *Col2a1* remains relatively stable and *Prg4* increases as joint development proceeds through cavitation and beyond^42–44^. In 2015, Spiers *et al.* applied the Illumina HumanMethylation450 array to DNA extracted from 179 human foetal brain samples between 3 and 26pcw^45^. Comparable to our dataset, they also identified that DNAm at ∼7% of captured CpGs significantly changed across the developmental window, despite the development within brain tissue of a more heterogenous cell population.

The largest hypermethylated dDMRs identified in this study predominantly map to transcriptional regulators of early embryogenesis and skeletal patterning, including *MEIS1, EN1, IRX1,* and *HOXA3.* Except for *HOXA3,* no correlations were identified between the expression of any of these genes and DNAm in the identified regions. However, the expression of these genes was low in all samples from 7pcw onwards and we postulate that we have captured a developmental window beyond the timeframe in which these genes impart their functional roles in cartilage development. Here, we appear to be observing hypermethylation of DNA as these gene regions become repressed, potentially to reinforce chromatin condensation. This is supported by the results of the GO analysis, which are enriched for a plethora of biological process terms relating to embryogenesis and skeletal development. Furthermore, the hypermethylated dDMRs were overrepresented in repressed regions of mature chondrocytes (36%, compared to 30% of all dDMRs) whereas none overlapped with gene transcriptional start sites. These observations also align with functional studies in murine models of limb development and the recent spatial maps of gene expression in human limb development^33^. *Meis1* is a regulator of proximodistal identity^46,47^ and was further shown by Zhang *et al* to be expressed specifically in the proximal hindlimb (developing femur) at 5.6pcw^33^. *En1* and *Irx1,* which orchestrate early murine limb bud patterning and digit formation, respectively are most highly expressed earlier and in the most distal locations in the limb^48^.

Conversely, several large hypomethylated regions mapped to genes encoding ECM proteins including Spondin-2 and Tenascin-X, highlighting the role of the DNA methylome in underlying transcriptional changes. We investigated the expression of the three genes mapping to the largest significant hypomethylated dDMRs, identifying that all significantly increased with decreasing DNAm. Across the developmental window, hypomethylated dDMRs were enriched in CREs, particularly annotated chondrocyte enhancers (29% overlapped with enhancers, compared to 25% of all dDMRs). This is consistent with earlier reports using an MSC *in vitro* model that, throughout differentiation, hypomethylation of DNA is enriched in enhancer regions^49^.

Dimorphism in DNAm levels between the biological sexes has been identified in a multitude of tissues across the human lifespan and is believed to contribute to the discrepancy in disease trajectories between males and females^50^. A recent study demonstrated that developmental sexual dimorphism in DNAm is tissue-specific, with a trend for hypomethylation in the placental tissues of female premature neonates, and hypermethylation in blood^37^. Due to the known discrepancies in the prevalence of knee OA in men and women, we investigated sDMRs at autosomal loci to look for sex-specific differences within the developing knee joint. The most significant sDMR mapped to the gene *NAB1*, encoding nerve growth factor induced (NGFI)-A-binding protein 1, a transcriptional repressor^51^ which is also the most significant DMP identified in neonatal blood. We found no significant interactions in our samples between sex and developmental stage upon DNAm. Sex differences in methylation profiles could potentially be reflective of different cell type populations in the same tissues, or subsets of articular chondrocytes between sexes. However, it is more likely that a yet-unknown biological impact is resulting in the observed sexual dimorphism, independent of cellular context, as many of the identified loci have also been identified in neonatal blood^37^.

Finally, we investigated the presence of epigenome-wide mQTLs within our foetal cartilage samples, discovering ∼400,000 significant SNV-CpG associations. Our co-localisation analysis revealed a shared impact of SNVs on DNAm and OA genetic risk. This confirms and builds upon our earlier report that the genetic and epigenetic interplay underlying OA risk can operate within chondrocytes from the start of life^21^. The majority (73%) of the co-localisations had not previously been identified in studies of aged cartilage, supporting our earlier hypothesis that some epigenetic mechanisms identified in the foetal samples may operate throughout the life course, whilst others may be constrained within the developmental period. Our enrichment analysis also demonstrated that the CpGs which are stringently regulated by SNVs in a tissue specific manner from the very start of life, are less amenable to modulation by their environment, highlighting their ability to differentially regulate a target gene throughout the life course. Whilst the genes mapping to the mQTL loci remain to be functionally validated, our *in-silico* analysis reveals putative regulatory mechanisms.

We identified novel OA-mQTLs mapping to the 3’UTR (2 CpGs) and downstream (1 CpG) of *CHST3,* within and adjacent to open chromatin regions. *CHST3* codes for chondroitin-6-*O*-sulfotransferase 1, an enzyme responsible for the sulfation of extracellular matrix (ECM) glycosaminoglycans (GAGs)^52^. Sulfation profiles of ECM GAGs within cartilage change throughout development and ageing and play a role in cell differentiation and tissue morphogenesis^52^. Mutations in *CHST3* lead to skeletal dysplasia, highlighting the important function of this protein in skeletogenesis^53^. Any conferred changes to the expression of the gene and its encoded protein, mediated through differential methylation, could potentially confer a subtle phenotypic shift within cartilage, decreasing its integrity to withstand mechanical forces over the life course.

We further identified OA-mQTLs in foetal cartilage that map to the genes *ALDH1A2* and *GDF5,* loci at which such effects have previously been identified in aged cartilage. On chromosome 15, two OA-mQTLs were identified falling within the first intron on *ALDH1A2,* encoding RALDH2, integral for all-*trans* retinoic acid (atRA) synthesis*. G*enetic studies of hand OA have been conducted on *ALDH1A2* based on the lead variant rs11071366, first identified in the Icelandic population^54^. Allelic expression imbalance at the locus showed a decrease in gene expression driven by the OA risk allele, which was present in osteochondral tissue from the trapezium, along with cartilage from the hip and knee^55,56^. This effect was also replicated in our targeted analysis of human foetal cartilage^21^. Furthermore, an OA-mQTL was identified at cg12031962, where the minor and effect allele, C, correlated with a decrease in DNAm^11^. In this study, we colocalised the OA risk SNV rs11071366 (A>T)^31^, which is in moderate pairwise linkage disequilibrium in European populations (r^2^=0.56). In our analysis of the foetal tissues, we also identified an OA-mQTL at the same site, where the effect allele, A, was also associated with decreased DNAm. Recent studies have shown that atRA has an anti-inflammatory effect within the joint, and that pharmacologically blocking its cellular metabolism exhibited a chondroprotective effect^56^. As an example of an epigenetic OA risk mechanism occurring throughout the life course, this knowledge could influence the key therapeutic window for future targeted interventions.

Due to the nature of the samples used in this study, it was not possible to use a replication cohort to validate our findings. One further limitation to this analysis is that it has not been possible to directly compare our findings to those in adult cartilage samples, nor studies of foetal methylation in blood and brain tissues due to batch effects and differences in the applied array technology. We hypothesis that such analyses would potentially reveal OA-mQTLs which become functionally manifest in later life, as opposed to exerting developmental, or lifelong functional effects.

In summary, we report the first comprehensive analysis of DNA methylation during human skeletal development. Our findings complement existing evidence that genetic factors linked to OA risk exert regulatory influences during the prenatal stage, underscoring the significance of early developmental processes in joint homeostasis and disease in later life. Moreover, our study showcases the effectiveness of mQTL mapping in pinpointing potential causal genetic sites associated with complex diseases across complex genomic regions. Future studies including both aged and developmental samples will identify potential subsets of genetic risk loci which may be active across the life course, requiring earlier interventions to prevent the onset and progression of disease. Our data supports our hypothesis of three categories of functional OA risk: 1) loci active only during development; 2) loci active throughout the life-course; 3) loci active only in mature/aged tissue. When this is known for a locus/pathway, intelligent timing of a pharmacological intervention can then be undertaken, and those which act in later life can be prioritised. Investigations that directly compare multiple joint tissues have also recently proven insightful into the molecular basis of the disease^8,57^. Such studies undertaken throughout the life course will be vital to further elucidate the tissue-specific gene regulatory networks contributing to disease, paving the way for therapeutic intervention.

## Acknowledgements

SJR is funded by The Royal Society (RGS\R1\231319), the JGW Patterson Foundation, the MRC-Versus Arthritis Centre for Integrated Research into Musculoskeletal Ageing (CIMA; MR/P020941/1 and MR/R502182/1), and a Versus Arthritis Career Development Fellowship (22615). DR is funded by NIH K99AR078352. AKS received research fellowship funding from the Wellcome Trust, Royal College of Surgeons of England, and the UK-US Fulbright commission.

## Author Contributions

Conceptualization, SJR; Methodology, EM, JS, AKS, SJR; Formal Analysis, EM, JS; Investigation, EM, JS, SEO, MJB, NW, LM, LO, AKS, SJR; Resources, NW, LM, LO; Data Curation, EM, JS, SJR; Writing-Original Draft, SJR; Writing-Reviewing and Editing; EM, JS, DAY, SJR; Visualization, SEO, SJR; Supervision, JS, SJR, DAY; Project administration, SJR; Funding Acquisition, SJR.

## Data Availability

Code used during this project are available on https://github.com/CBFLivUni/Epigenetics-OA-Risk-Human-Skeletal-Dev. Processed DNA methylation data for the 72 samples have been uploaded to the Gene Expression Omnibus (GEO) and will be made available on the publication of this article.

## Supplemental information

Supplementary Figures: Figures S1–S5

Supplementary Tables: Tables S1-14

Supplementary Methods: Additional methodological data which is too long to fit in the main body of the text. Contains the majority of the information regarding the bioinformatic analyses.

