## Supplementary material for "Epigenetic mechanisms of osteoarthritis risk in human skeletal development": Table S

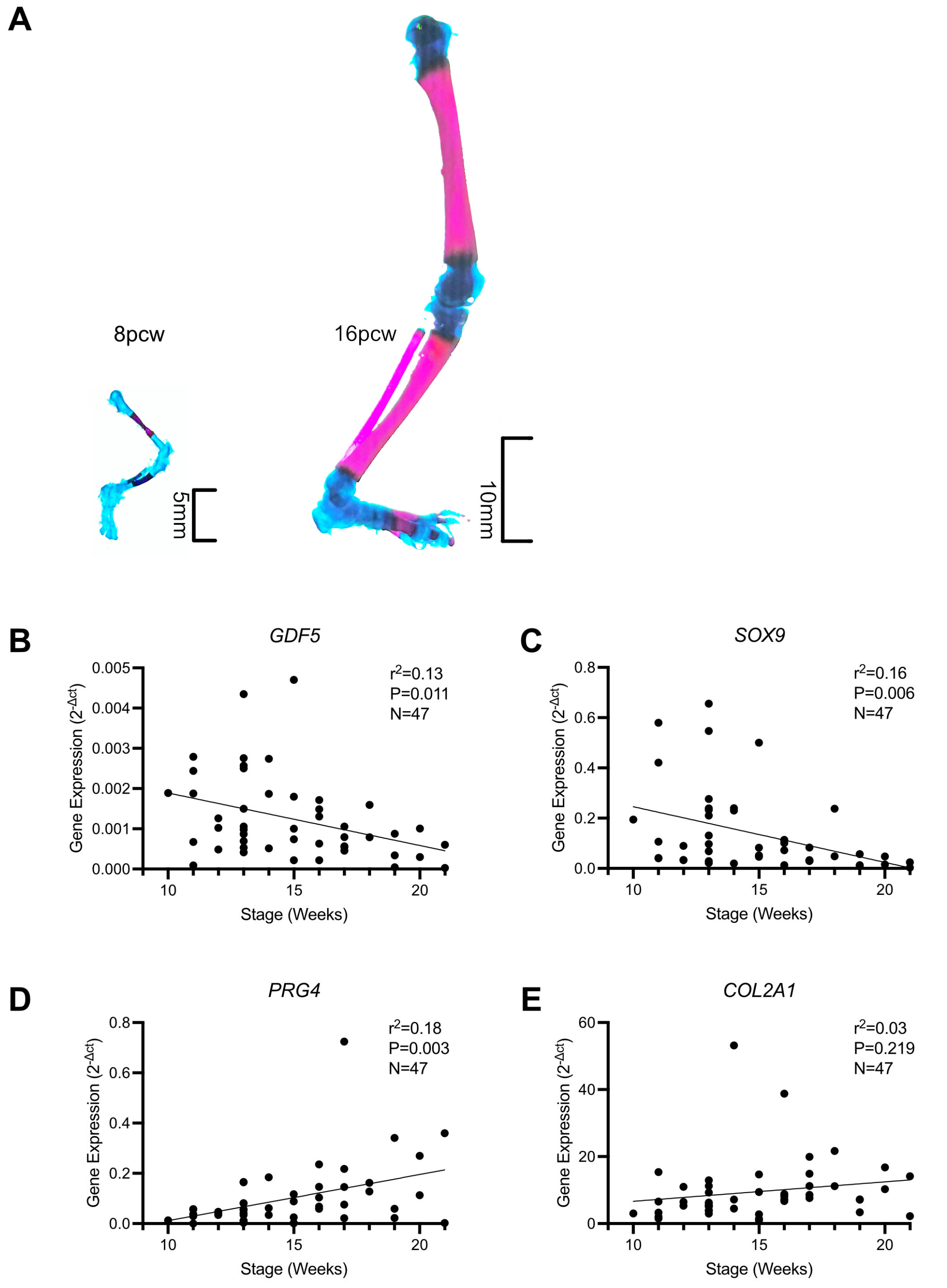


**Supplementary Figure 1. Characterisation of the human knee cartilage samples. A,** Alician blue and alizarin red staining of human foetal femur at 8pcw (left) and 16 pcw (right). **B-E,** Gene expression of cartilage progenitor (*GDF5* and *SOX9*) and chondrocyte (*PRG4* and *COL2A1*) makers in the samples. Simple linear regression was performed by the developmental stage.

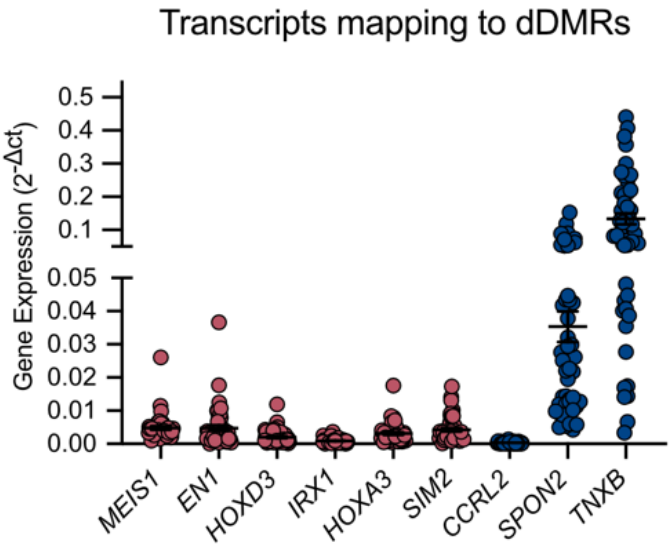


**Supplementary Figure 2.** Expression of genes mapping to the top hypermethylated (red) and hypomethylated (blue) developmental DMRs (dDMRs).

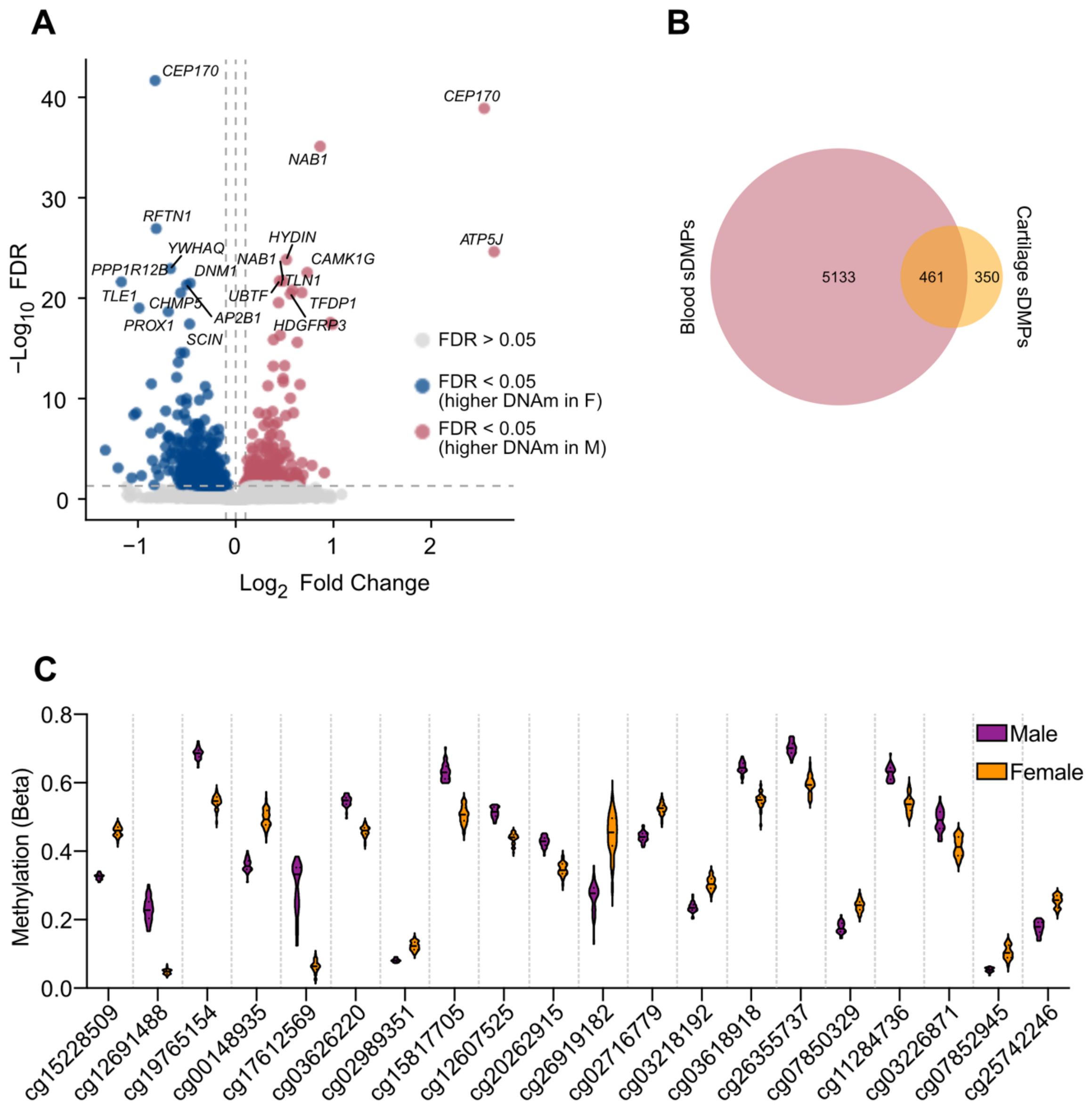


**Supplementary Figure 3. Sexual dimorphism in DNA methylation within human developmental cartilage. A,** Volcano plot of sDMPs. Grey, non-significant sDMPs (FDR adjusted P-value (P.adj) <0.05); Blue, significant sDMPs with higher methylation in female (F) samples (P.adj < 0.05); Red, significant sDMPs with higher methylation in male (M) samples (P.adj < 0.05). **B,** Venn diagram of overlap between sDMPs in neonatal blood reported in Santos *et al.* (pink) and foetal cartilage (orange). **C,** Methylation beta values in human foetal cartilage samples at the most significant sDMP CpGs. Purple, male; orange, female.

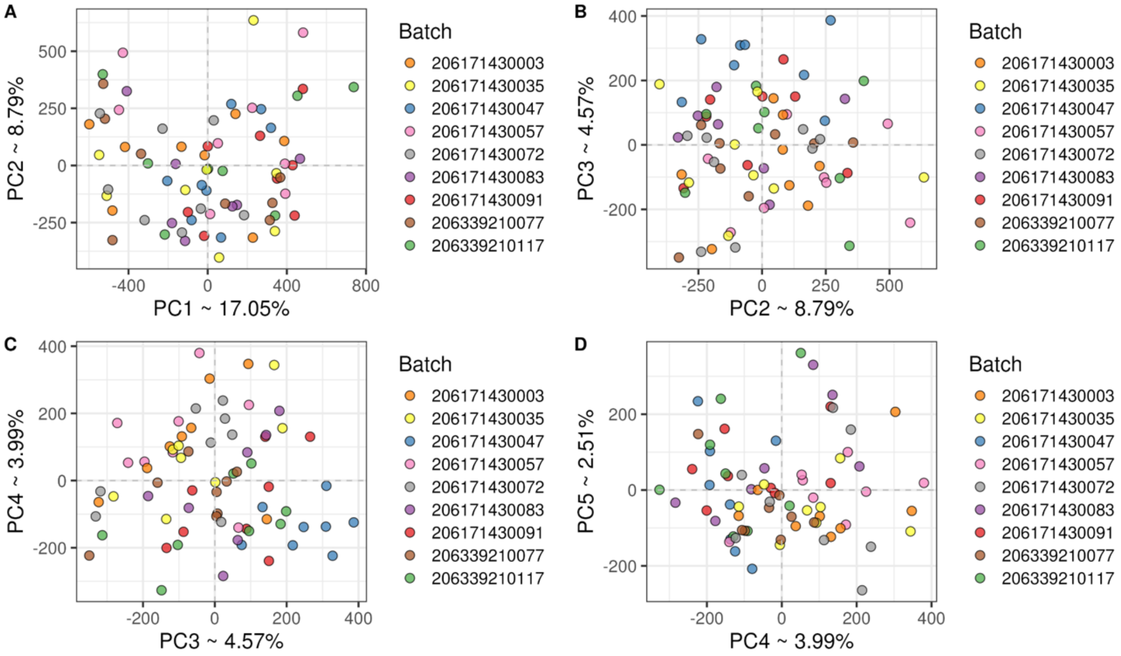


**Supplementary Figure 4. Assessment of batch effects by principal component analysis of the 71 foetal cartilage samples used in this study.** Principal component analysis of normalised DNAm samples coloured by Sentrix ID shows evidence of weak batch effects, observable in-association with PC3 and PC4.

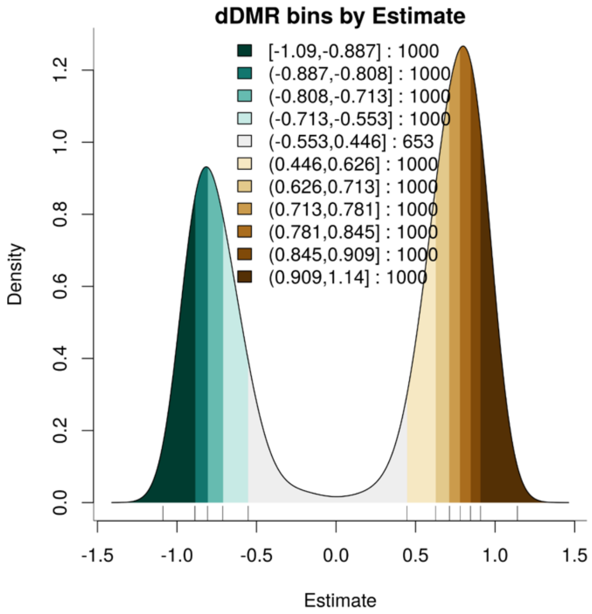


**Supplementary Figure 5. Developmental differentially methylated region (dDMR) bins for transcription factor motif analysis.** Histogram of the equally sized groups of developmental DMRs binned by Estimate (log2 fold change) that were used for transcription factor motif analysis with monaLisa.
