## Supplementary Methods for "Epigenetic mechanisms of osteoarthritis risk in human skeletal development"

### Sample isolation and nucleic acid extraction

Nascent articular and epiphyseal cartilage tissue (20-30mg) was taken from the distal end of the developing femur. As anticipated, adipose tissue was visible within the joint from ~19pcw and was thoroughly removed before cartilage isolation. In all cases, tissue was homogenised in 2ml screw-cap tubes containing 600μl RTL lysis buffer (AllPrep, Qiagen) at 2400rpm for 2.5min in 2mm Zirconia beads using the Mini BeadBeater 24 (both from BioSpec Products, USA). Samples were centrifuged at 13,000rpm for 30s before further homogenising at 2400rpm for 5min. Samples were centrifuged again before clarification through 0.2μM nylon filters (GeneFlow, UK). DNA was isolated using AllPrep (Qiagen) and RNA was isolated using Norgen DNA/RNA mini purification kit (Norgen BioTek Corp, Canada).

### Staining of bone and cartilage tissues

Foetal samples were stained following a previously published protocol[1], adapted for human tissue. Briefly, excess tissue was removed from the limb before fixing in 70% ethanol for 24hrs, then 95% ethanol for 24hrs. Ethanol was replaced with 0.03% Alcian blue solution for 1-3 days, depending on foetal stage, then washed with 95% ethanol for 6 hours, before replacement with 2% KOH solution for 12-24 hours. The KOH was changed to 0.005% Alizarin red for 12-24 hours, before clearance in 1%KOH:20% glycerol solution for 1-5 days. All stages took place with gentle agitation. The samples were imaged using brightfield optics.

### DNA methylation quality control

The R package minfi[2] (v1.4.6) was used to read in, process and perform quality control checks on methylation microarray data[3]. Bisulfite conversion and sex prediction checks were performed, and one sample was excluded from the analysis due to a mismatch between predicted and labelled sex. Non-autosomal sites were then excluded. Next, undetected probes (sum of Beta values across all samples equal to 0) and probes detected in less than 3 replicate beads were removed. Probes were then filtered to retain those with a detection p-value < 0.001 and any probes with missing data (marked as not applicable, NA) in any samples were excluded. Quantile normalisation was then applied to the microarray data. Finally, probes covered by known single nucleotide variants (SNVs) were removed using minfi and cross-reactive probes were removed using the R package maxprobes[3]. After quality control 678,267 probes representing CpG sites were retained for analysis.

### DNA methylation exploratory data analysis

Methylation data were transformed to M-values and filtered to remove the bottom 5% of probes by variance. Principal component analysis (PCA) was performed using the factoExtra R package (v1.0.7), using Z-scaled data as input (Fig.S4). Visual assessment of batch effect by PCA using Sentrix ID indicated PC3 and PC4 were associated with batch. Sentrix ID was therefore incorporated as a covariate in downstream statistical models.

### Differential methylation and differential methylated region analyses

Prior to statistical analyses, all data were transformed to M values. Probe-level differential methylation analysis was performed using the limma R package[4] (v3.55.5), using both the standard hypothesis testing method (null hypothesis of log2FC equal to 0) and the "treat" approach (null hypothesis of log2FC equal to or less than 0.1). The treat method of hypothesis testing was taken forward for stage-associated differential methylation analysis, while the standard hypothesis testing method was used for sex-associated differential methylation. Limma linear models included maternal BMI, maternal age and developmental stage as Z-scaled covariates and sex as a factor. Post-model fitting and testing, sex- and stage-DMPs were defined by filtration at an FDR of 0.001 and 0.05, respectively.

Differential methylated region (DMR) analysis with respect to the developmental stage was performed using the dmrff R package (v1.1.0) on the output p-values and standard errors of the probe-level differential methylation analysis. Probes were tested for association into a DMR with a maximum gap between probes of 1000 bp and DMRs defined as containing at least 2 probes and comprising the region between the 5' and 3'-most constituent probes. Resultant candidate sex DMRs (sDMRs) and developmental DMRs (dDMRs) were filtered by an FDR of 0.01 and 0.001, respectively.

**DMP clustering with mFuzz**

M values for significant stage DMPs were clustered using the Mfuzz R package[5] (v2.62.0) with the optimal cluster number chosen as 3, based on the elbow method. Minimum cluster membership was chosen as 0.99 with a seed value of 5 and the "fuzzifier" parameter was calculated as 1.15 using the "mestimate" function of Mfuzz.

**DMR overlap analysis**

DMR regions were overlapped within the bounds of Assay for Transposase Accessible Chromatin (ATAC)-Seq peaks previously generated in 12pcw foetal knee (distal femoral) cartilage[6] and the 15 chromatin-state model predictions for human cultured chondrocytes (E049) downloaded from the NIH ROADMAP Epigenomics Mapping Consortium[7]. Prior to overlap, the 15 ROADMAP states were collapsed into 5 functional classifications: enhancer (Enh, EnhG, EnhBiv), repressed (ZNF/Rpts, Quies, Het, ReprPC, ReprPCWk), transcription start site (TssBiv, TssA), flanking transcription start site (TssAFlnk, BivFlnk) and transcribed (TxWk, Tx, TxFlnk). Regions were overlapped using the intersect function in the bedtools (v2.31.1) package[8]. Presented frequencies and proportions represent overlaps between DMRs and ATAC regions/ROADMAP state classes, not the frequencies of unique members of either of these features. State enrichments were performed using two-sided Fisher's exact tests, testing for frequencies of overlaps with hypo- or hypermethylated DMRs.

**GO Meth term enrichment analysis.**

Gene ontology (GO) term enrichment analysis was performed on MFuzz clusters and sex DMPs using the gometh of the MissMethyl package[9] (v1.36.0), while DMR enrichment analysis made use of the goregion function. The Gene Ontology (GO) database was used with array.type="EPIC" argument provided to provide DMP-gene annotations. All 678,267 CpGs detected above thresholds were provided as background for all comparisons. Terms were called as significant with an FDR threshold < 0.05.

### TF Motif Enrichment Analysis

TF Motif PWMs were downloaded from HOCOMOCO v12[10] and filtered to retain motifs with a quality grade of C and above. MonaLisa (v1.9.0) was used to group all identified dDMRs into bins of 1000 DMRs by fold change[11]. To allow comparison between bins, the DMR sequences were trimmed to the median DMR size (253bp), centred around the centre of each DMR (Fig.S5). Enrichment statistics (Fisher's exact test) were calculated by comparing occurrences of each motif in a bin to its occurrences in all other bins. Hierarchical clustering of the motif similarity of the enriched motifs was used to visualise the enrichment scores.

### Genotype calling and genotype data filtration and quality control

Genotypes were called from raw idat files using gencall and associated array cluster and manifest files. The resultant gtc files were converted to vcf via the gtc_to_vcf.py script of the GTCtoVCF codebase (https://github.com/Illumina/GTCtoVCF). PLINK (v1.90b6.21) was used for subsequent analytic steps, except where otherwise stated[12]. Strand-flipped SNVs were identified by performing sample-wise merges with all remaining samples before all sample data was merged to one PLINK dataset. Genotype data filtering was performed using a minor allele frequency (MAF) > 0.01, while >0.975 was used as the variant and individual genotyping rate filtration threshold. X chromosome pseudoautosomal regions (PARs) were split to a separate chromosome, "26". Sex and interrelatedness checks were performed using PLINK.

### Imputation

Pre-imputation checks made use of the checkVCF utility (https://github.com/zhanxw/checkVCF/tree/master). All samples were submitted for imputation, while retaining only autosomes within each sample. Imputation was performed via submission to the Michigan Imputation Server (https://imputationserver.sph.umich.edu/ ) with population set to "EUR", genome build as "hg19" and using the haplotype reference consortium (HRC) as reference population[13,14]. Imputed vcf files were then filtered to split multiallelic sites into separate records and sites filtered by $r^2$>0.3. Imputed variants were then filtered with MAF > 0.05, alongside >0.975 for both variant and individual genotyping rates. Pre-imputation and post-imputation processing made use of bcftools (v1.16) and post-imputation checks utilised the ic utility (v1.0.9) of the batch McCarthy Group Tools suite[15]. After imputation and filtering, 5,394,299 variants were used in the subsequent mQTL analysis.

### 1000 genomes project PCA

PLINK2 format data (pgen, pvar and psam files) for the 1000 genomes project were converted to PLINK format data (.bed, .bim and .fam) and these were used as the reference population data[16]. For both reference and study data, A-T and C-G alleles were excluded and multiallelic sites were entirely excluded, using the "--biallelic strict" option. Next, sites of high linkage-disequilibrium (LD) were removed from the study data using a database of sites provided by the R package plinkQC (v0.3.4). Sites in the study data were then filtered with a 50kb sliding window, moving in 5kb steps and removing sites with a pairwise $r^2$>0.2. Sites in the reference data were then filtered by this pruned list of study data variants. Mismatched chromosome and position IDs were then identified and rectified, and variants orientated on the opposite strands between reference and study data were flipped, followed by the merging of the study and reference datasets. PCA was then performed using PLINK to obtain ancestry related principal components for inclusion in mQTL analysis.

1. Rigueur, D. & Lyons, K. M. Whole-mount skeletal staining. *Methods Mol Biol* **1130,** 113–121 (2014).
2. Aryee, M. J., Jaffe, A. E., Corrada-Bravo, H., Ladd-Acosta, C., Feinberg, A. P., Hansen, K. D. & Irizarry, R. A. Minfi: a flexible and comprehensive Bioconductor package for the analysis of Infinium DNA methylation microarrays. *Bioinformatics* **30,** 1363–1369 (2014).
3. Aryee, M. J., Jaffe, A. E., Corrada-Bravo, H., Ladd-Acosta, C., Feinberg, A. P., Hansen, K. D. & Irizarry, R. A. Minfi: a flexible and comprehensive Bioconductor package for the analysis of Infinium DNA methylation microarrays. *Bioinformatics* **30,** 1363–1369 (2014).
4. Ritchie, M. E., Phipson, B., Wu, D., Hu, Y., Law, C. W., Shi, W. & Smyth, G. K. limma powers differential expression analyses for RNA-sequencing and microarray studies. *Nucleic Acids Res* **43,** e47 (2015).
5. Kumar, L. & Futschik, M. E. Mfuzz: a software package for soft clustering of microarray data. *Bioinformation* **2,** 5–7 (2007).
6. Rice, S. J., Brumwell, A., Falk, J., Kehayova, Y. S., Casement, J., Parker, E., Hofer, I. M. J., Shepherd, C. & Loughlin, J. Genetic risk of osteoarthritis operates during human skeletogenesis. *Hum Mol Genet* (2022). doi:10.1093/HMG/DDAC251
7. Roadmap Epigenomics Consortium, Kundaje, A., Meuleman, W., Ernst, J., Bilenky, M., Yen, A., Heravi-Moussavi, A., Kheradpour, P., Zhang, Z., Wang, J., Ziller, M. J., Amin, V., Whitaker, J. W., Schultz, M. D., Ward, L. D., Sarkar, A., Quon, G., Sandstrom, R. S., Eaton, M. L., Wu, Y. C., Pfenning, A. R., Wang, X., Claussnitzer, M., Liu, Y., Coarfa, C., Harris, R. A., Shoresh, N., Epstein, C. B., Gjoneska, E., Leung, D., Xie, W., Hawkins, R. D., Lister, R., Hong, C., Gascard, P., Mungall, A. J., Moore, R., Chuah, E., Tam, A., Canfield, T. K., Hansen, R. S., Kaul, R., Sabo, P. J., Bansal, M. S., Carles, A., Dixon, J. R., Farh, K. H., Feizi, S., Karlic, R., Kim, A. R., Kulkarni, A., Li, D., Lowdon, R., Elliott, G., Mercer, T. R., Neph, S. J., Onuchic, V., Polak, P., Rajagopal, N., Ray, P., Sallari, R. C., Siebenthall, K. T., Sinnott-Armstrong, N. A., Stevens, M., Thurman, R. E., Wu, J., Zhang, B., Zhou, X., Beaudet, A. E., Boyer, L. A., De Jager, P. L., Farnham, P. J., Fisher, S. J., Haussler, D., Jones, S. J. M., Li, W., Marra, M. A., McManus, M. T., Sunyaev, S., Thomson, J. A., Tlsty, T. D., Tsai, L. H., Wang, W., Waterland, R. A., Zhang, M. Q., Chadwick, L. H., Bernstein, B. E., Costello, J. F., Ecker, J. R., Hirst, M., Meissner, A., Milosavljevic, A., Ren, B., Stamatoyannopoulos, J. A., Wang, T. & Kellis, M. Integrative analysis of 111 reference human epigenomes. *Nature* **518,** 317–329 (2015).
8. Quinlan, A. R. & Hall, I. M. BEDTools: a flexible suite of utilities for comparing genomic features. *Bioinformatics* **26,** 841–842 (2010).
9. Phipson, B., Maksimovic, J. & Oshlack, A. missMethyl: an R package for analyzing data from Illumina's HumanMethylation450 platform. *Bioinformatics* **32,** 286–288 (2016).
10. Vorontsov, I. E., Eliseeva, I. A., Zinkevich, A., Nikonov, M., Abramov, S., Boytsov, A., Kamenets, V., Kasianova, A., Kolmykov, S., Yevshin, I. S., Favorov, A., Medvedeva, Y. A., Jolma, A., Kolpakov, F., Makeev, V. J. & Kulakovskiy, I. V. HOCOMOCO in 2024: a rebuild of the curated collection of binding models for human and mouse transcription factors. *Nucleic Acids Res* **52,** D154–D163 (2024).

11. Machlab, D., Burger, L., Soneson, C., Rijli, F. M., Schübeler, D. & Stadler, M. B. monaLisa: an R/Bioconductor package for identifying regulatory motifs. *Bioinformatics* **38,** 2624–2625 (2022).
12. Purcell, S., Neale, B., Todd-Brown, K., Thomas, L., Ferreira, M. A. R., Bender, D., Maller, J., Sklar, P., De Bakker, P. I. W., Daly, M. J. & Sham, P. C. PLINK: a tool set for whole-genome association and population-based linkage analyses. *Am J Hum Genet* **81,** 559–575 (2007).
13. Das, S., Forer, L., Schönherr, S., Sidore, C., Locke, A. E., Kwong, A., Vrieze, S. I., Chew, E. Y., Levy, S., McGue, M., Schlessinger, D., Stambolian, D., Loh, P. R., Iacono, W. G., Swaroop, A., Scott, L. J., Cucca, F., Kronenberg, F., Boehnke, M., Abecasis, G. R. & Fuchsberger, C. Next-generation genotype imputation service and methods. *Nat Genet* **48,** 1284–1287 (2016).
14. McCarthy, S., Das, S., Kretzschmar, W., Delaneau, O., Wood, A. R., Teumer, A., Kang, H. M., Fuchsberger, C., Danecek, P., Sharp, K., Luo, Y., Sidore, C., Kwong, A., Timpson, N., Koskinen, S., Vrieze, S., Scott, L. J., Zhang, H., Mahajan, A., Veldink, J., Peters, U., Pato, C., Van Duijn, C. M., Gillies, C. E., Gandin, I., Mezzavilla, M., Gilly, A., Cocca, M., Traglia, M., Angius, A., Barrett, J. C., Boomsma, D., Branham, K., Breen, G., Brummett, C. M., Busonero, F., Campbell, H., Chan, A., Chen, S., Chew, E., Collins, F. S., Corbin, L. J., Smith, G. D., Dedoussis, G., Dorr, M., Farmaki, A. E., Ferrucci, L., Forer, L., Fraser, R. M., Gabriel, S., Levy, S., Groop, L., Harrison, T., Hattersley, A., Holmen, O. L., Hveem, K., Kretzler, M., Lee, J. C., McGue, M., Meitinger, T., Melzer, D., Min, J. L., Mohlke, K. L., Vincent, J. B., Nauck, M., Nickerson, D., Palotie, A., Pato, M., Pirastu, N., McInnis, M., Richards, J. B., Sala, C., Salomaa, V., Schlessinger, D., Schoenherr, S., Slagboom, P. E., Small, K., Spector, T., Stambolian, D., Tuke, M., Tuomilehto, J., Van Den Berg, L. H., Van Rheenen, W., Volker, U., Wijmenga, C., Toniolo, D., Zeggini, E., Gasparini, P., Sampson, M. G., Wilson, J. F., Frayling, T., De Bakker, P. I. W., Swertz, M. A., McCarroll, S., Kooperberg, C., Dekker, A., Altshuler, D., Willer, C., Iacono, W., Ripatti, S., Soranzo, N., Walter, K., Swaroop, A., Cucca, F., Anderson, C. A., Myers, R. M., Boehnke, M., McCarthy, M. I., Durbin, R., Abecasis, G. & Marchini, J. A reference panel of 64,976 haplotypes for genotype imputation. *Nat Genet* **48,** 1279–1283 (2016).
15. Li, H. A statistical framework for SNP calling, mutation discovery, association mapping and population genetical parameter estimation from sequencing data. *Bioinformatics* **27,** 2987–2993 (2011).
16. Chang, C. C., Chow, C. C., Tellier, L. C. A. M., Vattikuti, S., Purcell, S. M. & Lee, J. J. Second-generation PLINK: rising to the challenge of larger and richer datasets. *Gigascience* **4,** (2015).
